# Reducing spatial clustering to prevent tuberculosis transmission in a busy Zambian hospital: a modelling study based on person movements, environmental and clinical data

**DOI:** 10.64898/2025.12.24.25342956

**Authors:** Nicolas Banholzer, Guy Muula, Fiona Mureithi, Esau Banda, Pascal Bittel, Lavinia Furrer, David Kronthaler, Remo Schmutz, Matthias Egger, Carolyn Bolton, Lukas Fenner

## Abstract

**Background:** Hospitals in high-burden tuberculosis (TB) settings are important sites of *Mycobacterium tuberculosis* (*Mtb)* transmission, yet the impact of infection prevention and control (IPC) measures targeting crowding is poorly understood. We assessed the effects of simple interventions to reduce spatial clustering and airborne transmission in a Zambian hospital.

**Methods and findings:** From June to August 2024, we prospectively collected clinical data on presumptive (symptom-based screening) and confirmed (Xpert-positive, chest X-ray) TB patients, indoor CO_2_ levels, bioaerosol samples, and continuous person movements (optical sensors) in the hospital’s main waiting hall. Airborne *Mtb* DNA was detected using hourly cyclonic bioaerosol sampling. Using a spatiotemporal Wells–Riley model integrating ventilation, proximity between visitors, and movement patterns, we estimated *Mtb* transmission risk under routine conditions and during two IPC interventions: (1) an optimised waiting-area layout with physical distancing measures; and (2) an added one-way patient flow system.

During 52 days, 668 presumptive and 45 confirmed TB patients visited the hospital, and 671 840 person movements were recorded. Despite excellent natural ventilation (median CO_2_ 455 ppm; 8.0 air changes per hour), airborne *Mtb* was detected on six days. The first intervention reduced spatial clustering by an estimated 24% (95% credible interval [CrI] 13–32) and the second by 13% (95% CrI 1–23). The interventions lowered *Mtb* transmission risk by an estimated 39% (95% CrI 29–48) and 21% (95% CrI 9–32), respectively. Over the four weeks of implementation, they collectively averted an estimated 16 (95% CrI 8–26) infections.

**Conclusions:** In this high-burden, well-ventilated hospital, short-range exposure remained a driver of airborne *Mtb* transmission. Simple, low-cost operational changes to reduce clustering substantially decreased proximity-driven transmission risk. Integrating proximity-focused strategies can meaningfully strengthen IPC in resource-limited healthcare settings.

## Introduction

Tuberculosis (TB), caused by *Mycobacterium tuberculosis* (*Mtb*), remains one of the leading causes of death from infectious disease worldwide, particularly in sub-Saharan Africa where TB and HIV epidemics intersect.^1^ Transmission of *Mtb* occurs through airborne particles released by individuals with pulmonary TB, and the infection risk depends on factors such as ventilation, duration of exposure and physical distance between people (proximity). Despite decades of control efforts, the mechanisms and settings driving transmission remain incompletely understood,^2^ partly because directly measuring airborne transmission is difficult.

Healthcare facilities attended by people with presumptive or confirmed TB represent high-risk environments for nosocomial transmission. Previous studies have assessed infection prevention and control (IPC) measures such as improved ventilation, mask use, and isolation of infectious patients, and have demonstrated their potential to reduce *Mtb* transmission.^3–7^ However, the effectiveness of operational interventions that reduce the spatial clustering of people remains unclear. Empirical data on how individuals move and interact in clinical spaces are rarely available, and most risk models assume well-mixed indoor air,^2,8,9^ overlooking short-range transmission gradients driven by proximity. Recent advances in environmental monitoring and indoor movement-tracking technologies now allow for detailed characterisation of spatial and temporal crowding patterns in hospitals, offering an opportunity to refine infection models and evaluate real-world IPC strategies.^7,10^

In this study, we conducted a longitudinal field study in a Zambian hospital with high TB/HIV-burden, integrating clinical, environmental, and person-tracking data to quantify the risk of airborne *Mtb* transmission. Using a spatiotemporal extension of the Wells–Riley model that incorporates proximity between people, we estimated the expected number of *Mtb* infections under routine conditions and during two interventions designed to reduce spatial clustering: an infrastructure modification coupled with physical distancing measures to increase spatial separation in the waiting area and reduce proximity; and an additional patient flow system to decrease the number of close-contact encounters.

## Methods

### Study setting and interventions

We collected clinical, environmental and person-tracking data for 52 days from June 17 to August 22, 2024, in the waiting hall of a hospital in Lusaka, Zambia (Figure 1). The hospital is located in a large settlement of formal and semi-formal housing where TB and HIV are highly prevalent. It provides tuberculosis, HIV, and general outpatient services continuously, operating 24 hours a day, seven days a week. We collected data during the busy hours from 6 am to 6 pm under three different study conditions (see Supplementary Table S1 for details): a baseline phase (24 days) with routine clinic conditions for nine days at the start and fifteen days at the end of the study; a first intervention phase (14 days) with a redesigned waiting area to achieve a more uniform spatial distribution of patients, as well as signage and reminders to encourage physical distancing; and a second intervention phase (14 days) that added a patient flow system to decrease the number of close-contact interactions.

**Figure 1:**
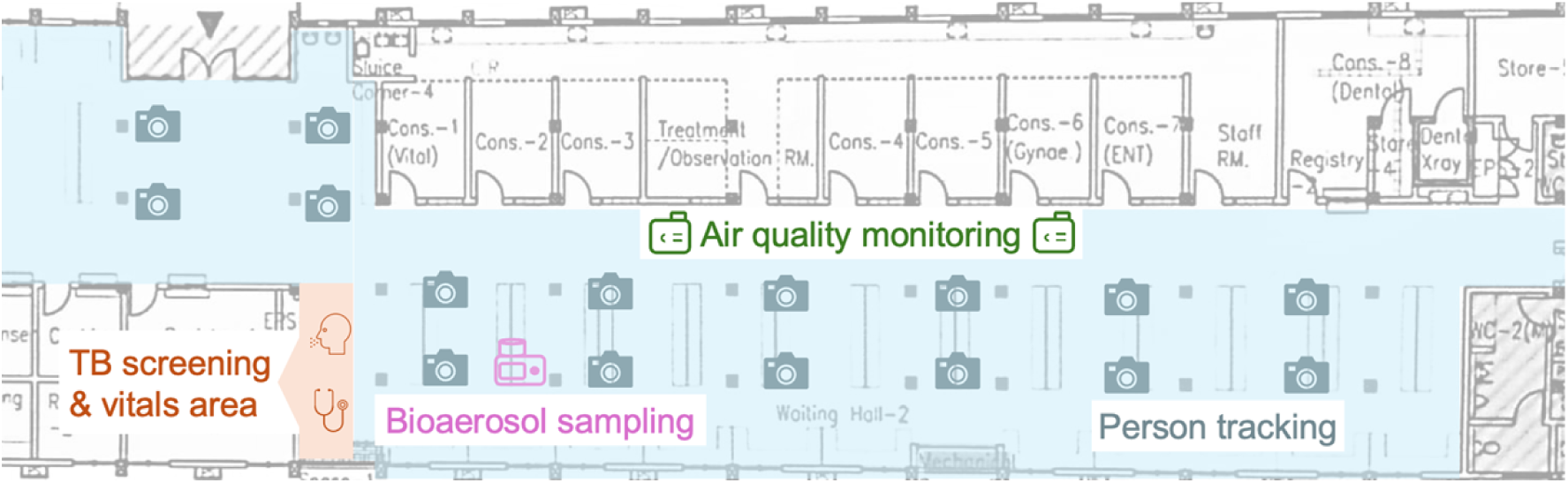
Schematic view of the study setting and interventions. Person movements were recorded via optical sensors in the main waiting area of the hospital (blue) and linked to TB patients via the time they presented in the TB screening area (orange). CO_2_ levels were recorded with air quality monitoring devices (green) and linked with person counts to compute daily outdoor air exchange rates. A cyclonic bioaerosol sampling device (pink) was run for ∼90 min per day to detect airborne *Mycobacterium tuberculosis*.

We aimed for a sample of 14 days per study phase to capture sufficient variation in crowding, ventilation and TB burden. Because patient volumes, ventilation patterns, and TB case burden can vary over time, we included baseline phases at both the beginning and end of the study to capture temporal trends in routine clinic conditions. Descriptive data are presented for all four phases (pre-intervention baseline, first and second intervention, post-intervention baseline) and the modelling approach employed for effect estimation was constructed to explicitly account for and remove observed seasonal trends, thereby mitigating potential bias.

### TB case definitions

All patients entering the hospital underwent symptom-based screening according to WHO guidelines (cough, fever, night sweats, or weight loss).^11^ For already diagnosed TB patients on treatment and people with presumptive TB, demographic and clinical characteristics (age, sex, self-reported HIV status, and vital signs) were recorded, and diagnostic investigations were performed according to the physicians’ decision, including Xpert Ultra testing on a single sputum sample and a chest X-ray examination when clinically indicated. People with confirmed TB (Xpert- or X-ray-positive) were considered infectious, except those who had already received anti-TB treatment for more than four weeks. People with presumptive TB and negative, invalid, or missing test results were considered potentially infectious, consistent with recent evidence showing that individuals with TB-like symptoms can generate aerosolised *Mtb* even in the absence of a confirmed diagnosis.^12,13^ Throughout the study, mask use was high among hospital staff but rare among attendees.

### Person-movement tracking

As in previous studies in a South African clinic,^7,10^ we used an anonymous person tracking system with optical sensors (Xovis, Zollikofen, Switzerland) to track person movements at 1-second intervals in the main waiting area of the Zambian hospital. The time-stamped tracking data consisted of a person’s position (x-y coordinates) and a unique person ID. The sensors could recognise the person’s gender and distinguish patients from healthcare workers, who wore a specific paper tag on their chest. Data was continuously transferred via a secure SFTP server to the Institute of Social and Preventive Medicine, University of Bern, for further analysis.

We included 52 days with uninterrupted tracking. Owing to technical issues, person-level attributes (gender and healthcare worker) were unavailable for the first ten study days. Person movements were linked with TB patients based on the time they presented in the TB screening area. We used an R Shiny app (Supplementary Figure S1) to identify and manually rejoin interrupted movements of TB patients to provide a more accurate representation of their spatiotemporal locations in the hospital (Supplementary Text A).

### Environmental monitoring

Two air quality monitors (Aranet4 Home, SAF Tehnika, Riga, Latvia) recorded indoor CO_2_ levels (parts per million) at 5-min intervals in the waiting area. Their measurements were highly correlated but slightly higher for the device in the more crowded part of the hospital (Supplementary Figure S2), which we chose for computation of outdoor air exchange rates to assess ventilation (Supplementary Text B).^14^

### Bioaerosol sampling

We collected airborne *Mtb* using a cyclonic bioaerosol sampling device (Coriolis Micro Air; Bertin Instruments, France), running at 300 L/min and collecting 15 mL of phosphate-buffered saline.^15^ The sampling device ran for 10 min each hour from 8am to 4pm (∼90 min per day). Samples were transported to the laboratory on the same day and stored at *−*80°C. At the end of the study, samples were concentrated by ultrafiltration and GeneXpert Ultra tests were performed.

### Quantifying spatial clustering

We examined spatial density in the waiting area by rasterising the space into a grid of *n* squared cells each with a length of 0.5m. To approximate close-proximity interactions, the person counts *p* were smoothed using a Gaussian kernel density estimator with a bandwidth of 1.5m. Spatial density was quantified with the Gini coefficient 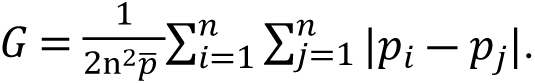. This coefficient compares the observed person counts with a uniform distribution, with larger values indicating higher clustering (larger deviation from a uniform spread of people).

### Modelling airborne *Mtb* transmission

Figure 2 shows a visual summary of our modelling and estimation approach; see Supplementary Text C for details and assumptions.

**Figure 2:**
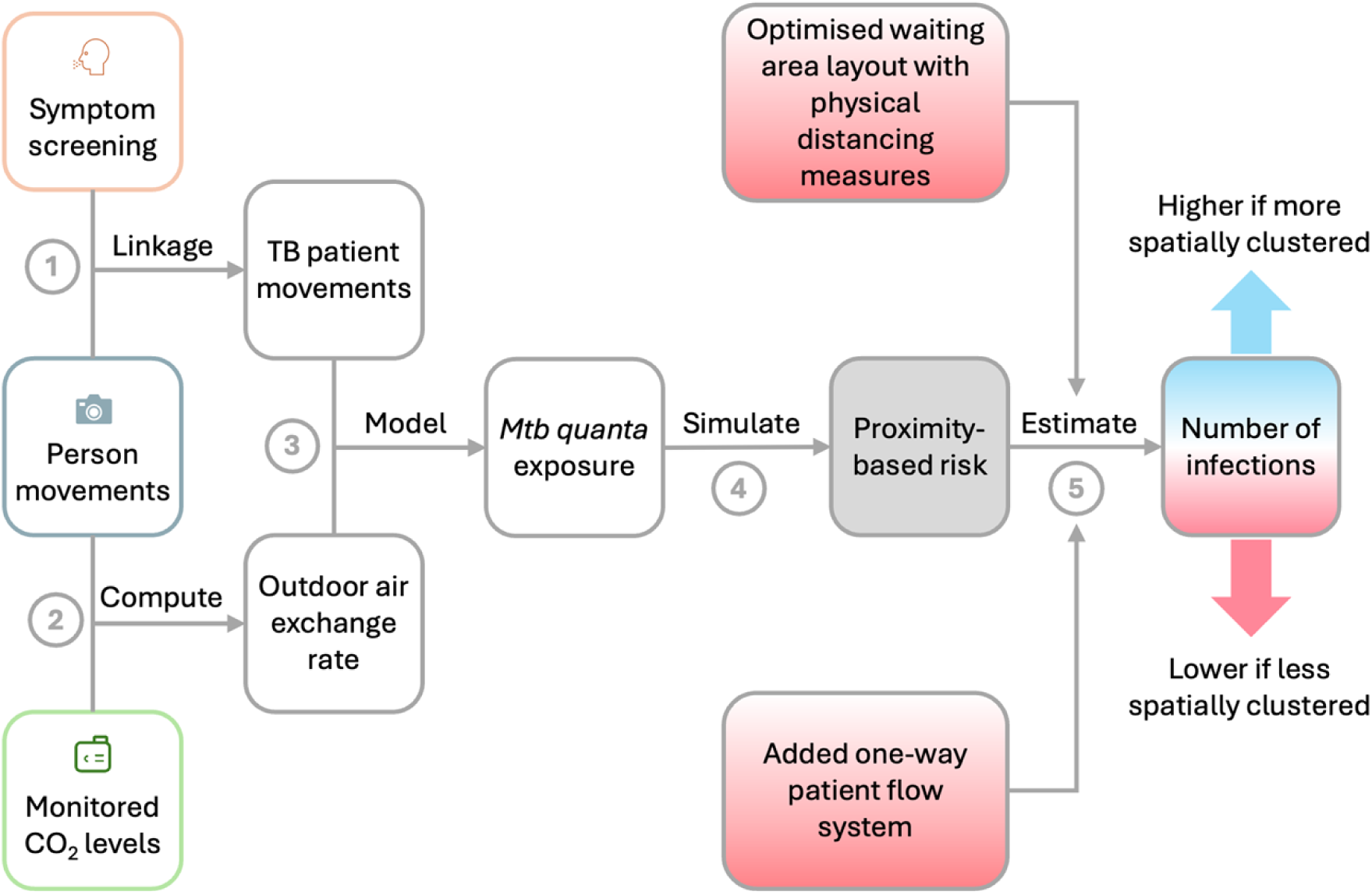
Visual summary of the modelling and estimation approach to assess intervention effects. (1) Person movements were linked with TB patients based on the time they presented in the area for TB symptom screening. (2) Person movements and CO_2_ levels were combined to compute the outdoor air exchange rate. (3) TB patient movements and daily outdoor air exchange rates were integrated into the spatiotemporal Wells-Riley model to estimate the quanta concentration in the waiting hall of the hospital. (4) *Mycobacterium tuberculosis* (*Mtb)* transmission risk was simulated with two model configurations, one considering proximity between hospital attendees (proximity-based infection risk) and one assuming a single, well-mixed airspace (uniform infection risk). (5) The ratio of the expected number of *Mtb* infections between both models (proximity-based/uniform) was computed per day and compared between study phases. An estimated decrease in the ratio would indicate that interventions reduced spatial clustering and the number of new *Mtb* infections.

We modelled *Mtb* transmission risk using a spatiotemporal extension of the Wells–Riley equation,^7^ which models airborne infection risk as a function of exposure to quanta (doses of infectious airborne particles).^2,8,9^ Quanta were generated by infectious individuals according to activity-specific emission rates and assumed to diffuse radially within the horizontal plane while remaining vertically well mixed. The local quanta concentration changed over time as a function of generation, diffusion, and removal (outdoor air exchange, bacterial inactivation, and gravitational settling). The infection risk was calculated for each person movement from its time-resolved exposure to local quanta concentrations and activity-specific breathing rates. The risks were aggregated to estimate the expected number of *Mtb* infections.

We performed 100 Monte Carlo simulations to consider uncertainty about quanta generation and removal rates. In each simulation, we assumed all people with confirmed TB were infectious. To account for potential infectiousness despite negative test results,^13^ we incorporated a 10–30% probability of infectiousness for people with presumptive TB, reflecting sensitivity of the Xpert Ultra test in routine clinical settings.^16^ We assumed that presumptive and confirmed TB patients were equally infectious. To examine the impact of this assumption, we re-estimated the expected number of *Mtb* infections under two alternative scenarios: (i) individuals with confirmed TB generated quanta at eight times the rate of those with presumptive TB;^17^ and (ii) individuals with HIV generated quanta at six times the rate of those without HIV.^18^

### Estimation of intervention effects on *Mtb* transmission

To evaluate intervention effects, we compared two model configurations: one where infection risk varied with spatial proximity to infectious individuals, and another assuming a well-mixed airspace with uniform risk. This approach allowed us to differentiate proximity-related transmission from overall transmission driven purely by the number of infectious individuals present. We quantified the intervention effect by the change in the ratio of infections between the two models (proximity-based/well-mixed), with a higher ratio indicating a larger contribution of proximity to transmission. Given that changes in crowding can influence proximity, we divided this ratio by total person-time to isolate the impact of interventions on transmission specifically via their effect on the spatial distribution of people.

The effects of interventions on spatial clustering and airborne transmission were estimated using Beta and log-linear regression models. Both models were adjusted for weekday effects to account for difficulties in enforcing interventions on crowded weekdays, as reported by our local study team. Bayesian models were estimated using Stan v2.32.2 with the brms R package v.2.20.0. Simulation results were summarised with medians and 95% quantiles and estimation results were pooled across simulations and reported with medians and 95% credible intervals (CrIs). Modelling and analyses were performed in Python v.3.13.0 and R v.4.4.2. Categorical variables were summarised with counts (proportion, %) and numerical variables with medians (interquartile ranges, IQRs).

### Ethics statement

We obtained approval from the local and the national ethics committees (UNZA-BREC 4763-2023) and from the National Health Research Authority (ref. no. NHRA-1116/11/04/2024). We obtained an informed consent waiver to collect a minimal dataset (including age, gender, TB/HIV information, diagnostic results) from people with presumptive or confirmed TB, since this information was collected anonymously as part of routine clinical procedures. In addition, the project was approved by the Cantonal Ethics Committee of Bern, Switzerland (reference no. PB_2016-00273).

## Results

### Clinical, environmental and person-tracking data

Table 1 shows a summary of the clinical, environmental and person tracking data. During 52 study days, 668 presumptive and 45 confirmed TB patients visited the hospital (see linelist in Supplementary Table S2): 313 female (44%) and 117 reported living with HIV (16%). Six TB patients were already on anti-TB treatment, four of them for less than a month, thus still considered infectious. While more people with confirmed TB visited the hospital towards the end of the study, the number of patients with presumptive TB steadily decreased over the study (Figure 3A). This trend may be partially attributed to seasonal effects, specifically the rise in ambient temperatures (Table 1), potentially leading to fewer hospital visits for cold and influenza-like symptoms.

**Figure 3:**
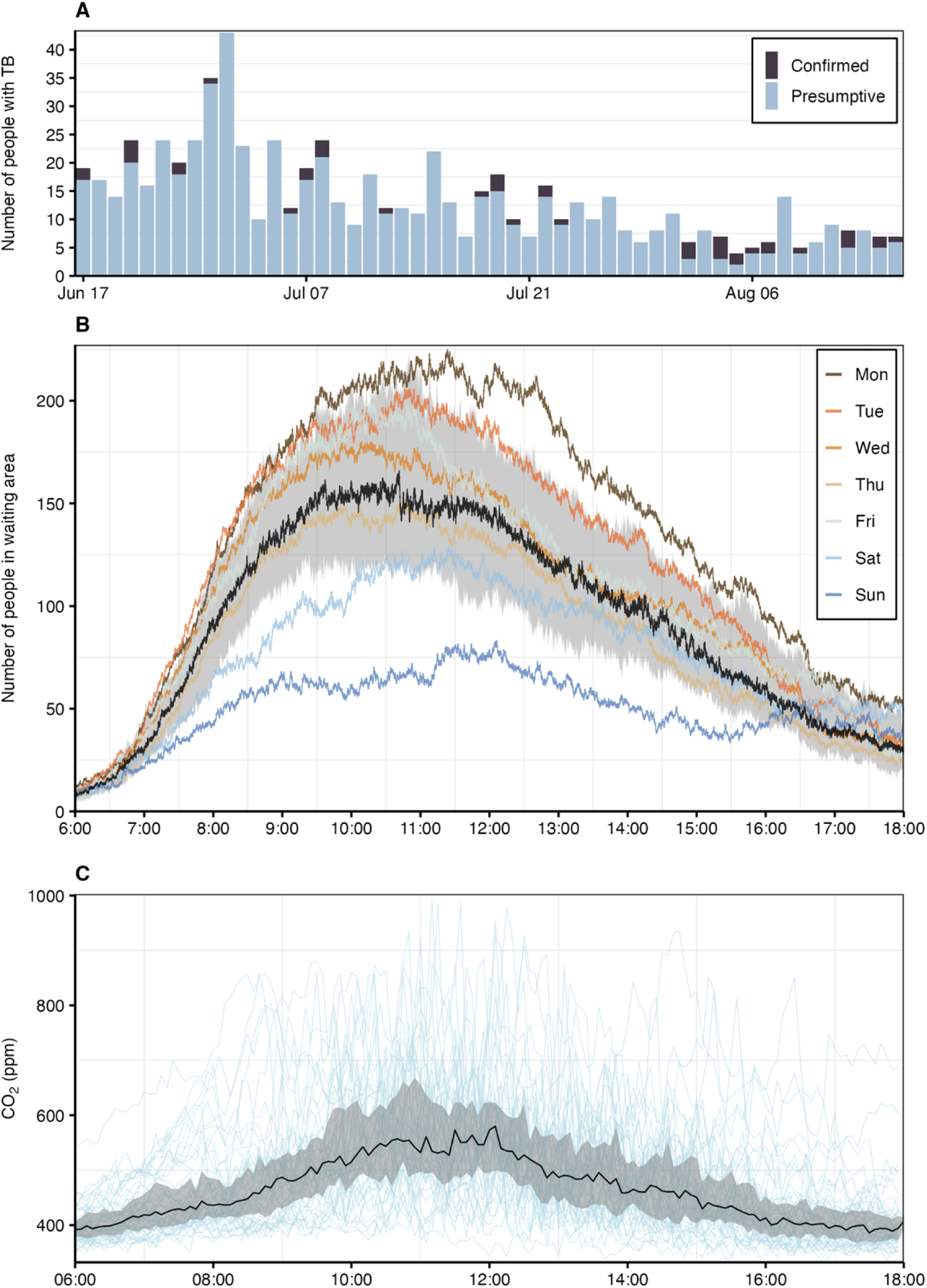
Clinical, environmental and person-tracking data. (**A)** Daily number of people with presumptive and confirmed TB. **(B)** Number of people in the hospital’s waiting area (median as black line, interquartile range as interval and weekday averages as coloured lines). **(C)** Intraday CO_2_ levels (median as black line, interquartile range as interval and individual study days as light blue lines).

**Table 1:**
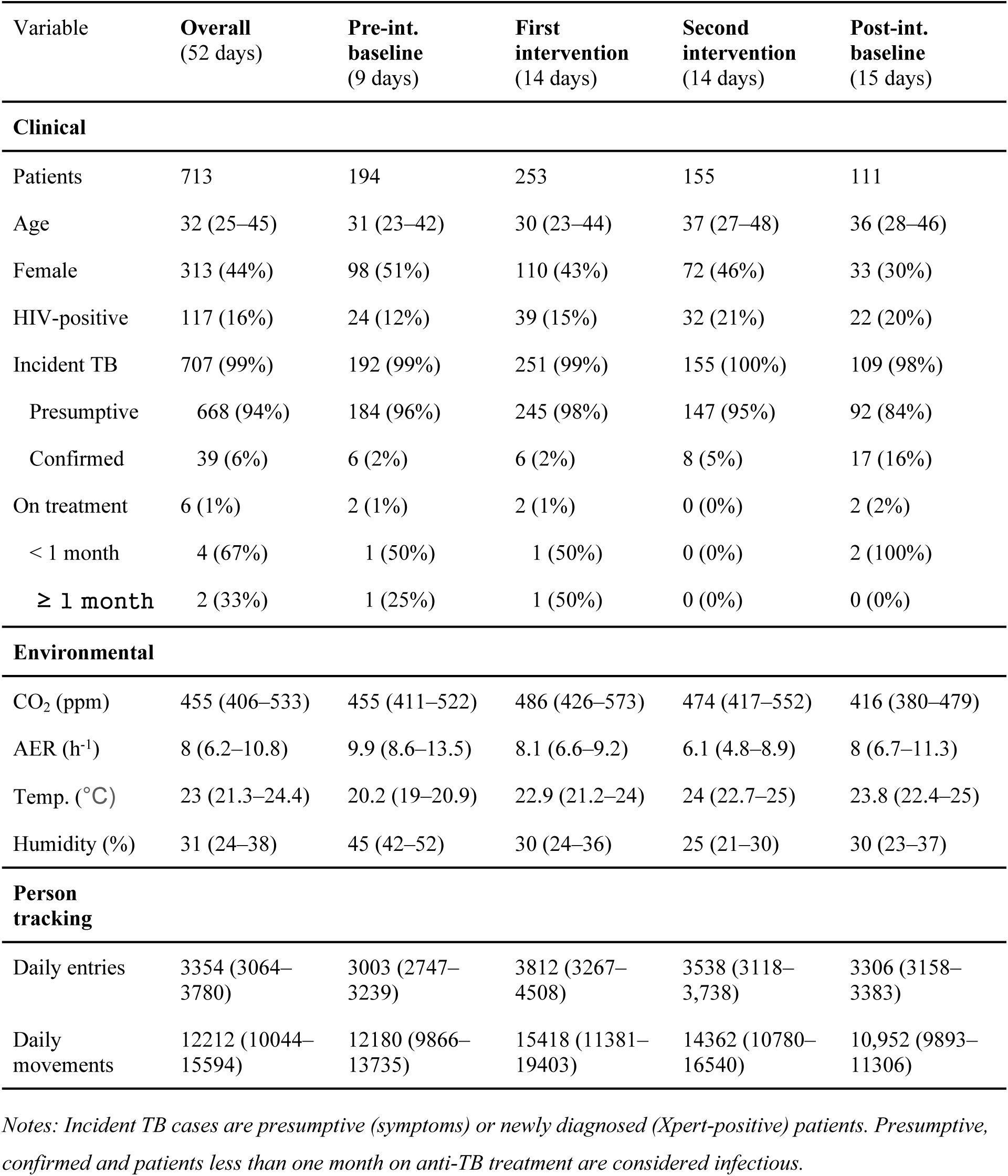
Study population and setting. Summary of clinical (patient characteristics, presumptive and confirmed TB cases), environmental (CO_2_, air exchange rate [AER], temperature and relative humidity), and person-tracking (recorded entries and person movements) data overall and by study phase. Categorical variables are shown with counts (proportion in %) and continuous variables with median (interquartile range). Detailed clinical linelist data is proved in Supplementary Table S1.

Optical sensors recorded 178,510 entries into the hospital and 671,840 person movements. The daily number of entries and person movements varied between study phases and exhibited a decreasing trend after the pre-intervention baseline phase (Table 1). The median duration per movement was 1.3 min (IQR 0.5–4.7) and the distribution was right-skewed (Supplementary Figure S3), reflecting a higher proportion of transient movements. The number of people in the waiting area peaked at 188 (IQR 160–242) before noon. On average, occupancy was lower on weekends and higher on weekdays such as Mondays and Tuesdays (Figure 3B).

Indoor CO_2_ levels typically remained below 600 ppm (Figure 3C), with a median of 455 ppm (IQR 406–533). These low concentrations support effective natural ventilation, with a median daily air exchange rate of 8.0 h−1 (IQR 6.2–10.8) (Supplementary Figure S4). Despite high outdoor air exchange, *Mtb* was detected in bioaerosols on six study days (Supplementary Table S3), mainly during the earlier phase of the study, with a median cycle threshold (Ct) value of 28.6 (IQR 27.3–31.6).

### Spatial clustering of people by study phase

The spatial clustering of people, measured using the Gini coefficient, increased as the waiting area became more crowded (Supplementary Figure S5). The daily median Gini coefficient was 0.55 (IQR 0.51–0.58) during baseline, 0.51 (IQR 0.45–0.55) during the first intervention with the optimised waiting area layout and physical distancing measures, and 0.52 (IQR 0.48–0.62) during the second intervention with the added patient flow system (Supplementary Figure S6). Adjusted for weekday effects, the Gini coefficient decreased by 24% (95% credible interval [CrI] 13–32) during the first and 13% (95% CrI 1–23) during the second intervention (Figure 4A), suggesting that spatial clustering was lower during both interventions.

**Figure 4:**
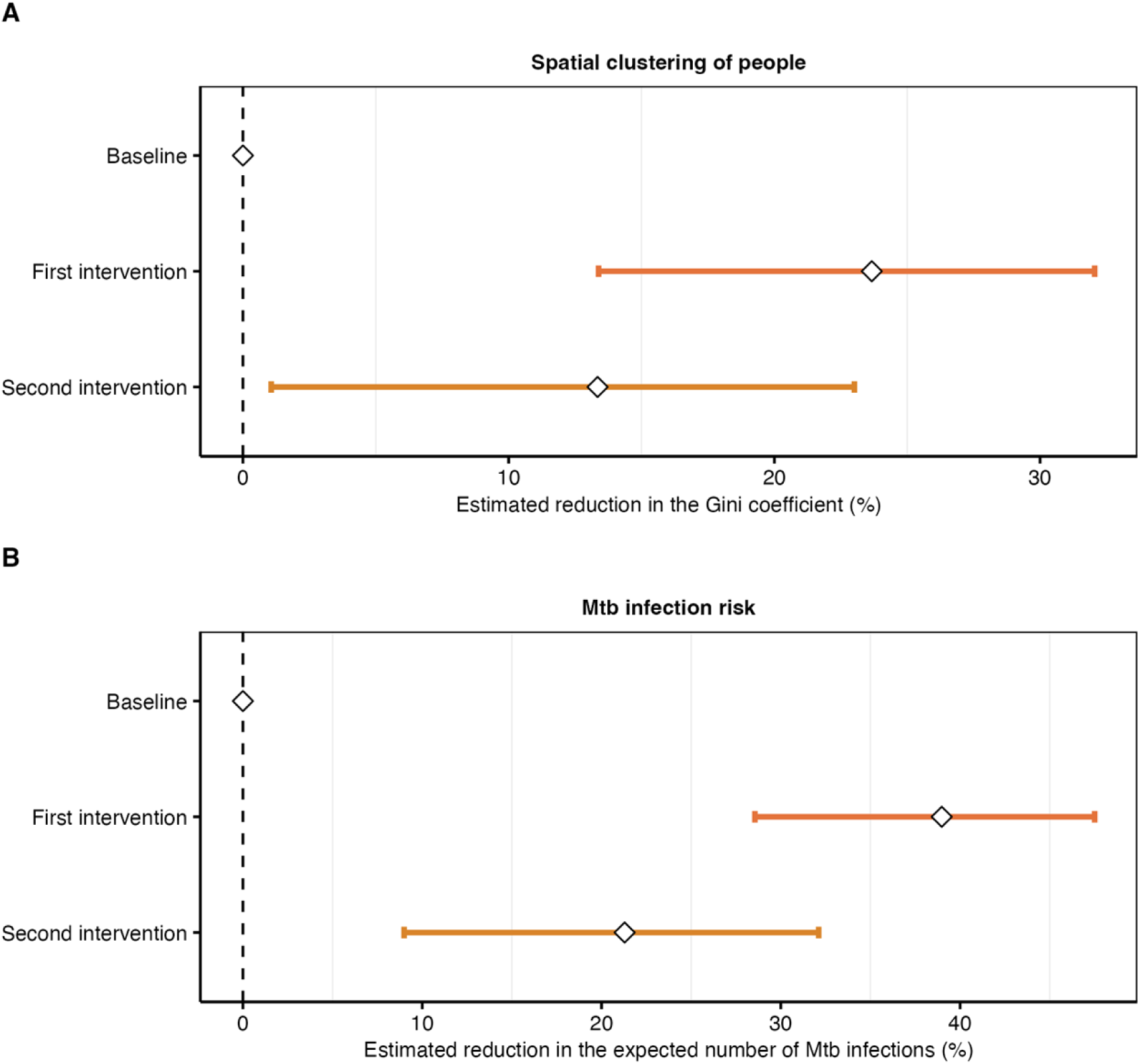
Effects of proximity-targeting interventions on spatial clustering and *Mtb* transmission. **(A)** Estimated reduction in the Gini coefficient, which was used to measure the spatial clustering of people in the hospital’s waiting area. **(B)** Estimated reduction in the expected number of *Mtb* infections. Median reductions as dots and 95% credible intervals (CrIs) as lines. Baseline: routine clinic conditions; first intervention: optimised waiting area layout and guided physical distancing; second intervention: optimised waiting area layout, guided physical distancing and patient flow system.

### *Mtb* infection risk and effects of interventions

Based on our model, we expected 7 (95% quantile 3–32) *Mtb* infections to occur in the hospital waiting area over the 52-day study period. The modelled infection risk was sensitive to assumptions about infectiousness: if people with confirmed TB or people with HIV were assumed more infectious, the expected number of infections would have been 82% (95% quantile 13–170) or 38% (95% quantile 6–126) higher, respectively (Supplementary Figure S7). The rate of infection tended to be lower for female than male hospital attendees (–18%, 95% quantile –50–19) (Supplementary Figure S8). While the rate was similar between healthcare workers and other hospital attendees (–2%, 95% quantile –55–56), healthcare workers probably had a lower risk because of consistent mask use, which was not modelled.

The modelled daily expected number of *Mtb* infections showed a decreasing trend over the study period (Supplementary Figure S9), coinciding with reductions in the number of presumptive TB patients (Figure 3A) and changes in hospital entries and person movements (Table 1). To isolate the impact of proximity-targeting interventions from temporal variation in TB burden and occupancy, we modelled the ratio of *Mtb* infections as estimated by the proximity-based model to those estimated by a well-mixed model, and normalised this ratio by total person-time. This ratio and normalisation effectively removed the temporal trend from the outcome (Supplementary Figure S10) and confirmed that proximity contributed to transmission risk (Supplementary Figure S11).

Adjusted for weekday effects, *Mtb* transmission was estimated to be 39% (95% CrI 29–48) lower during the first and 21% (95% CrI 9–32) lower during the second intervention (Figure 4B). Extrapolating from the estimated effects, there would have been 11 (95% CrI 5–18) and 5 (95% CrI 2–9) more infections during the first and second two-week intervention phase, respectively, if the interventions had not been implemented. Collectively, the interventions likely prevented 16 (95% CrI 8–26) *Mtb* infections during their four weeks of implementation.

## Discussion

In this longitudinal field study conducted in a busy primary care hospital in Lusaka, Zambia, we quantified airborne *Mtb* transmission risk using clinical, environmental, and high-resolution person-movement data. Despite excellent natural ventilation, airborne *Mtb* was intermittently detected in the waiting hall, and our spatiotemporal Wells–Riley model estimated that short-range exposure remained an important driver of transmission. Simple operational interventions reduced spatial clustering and lowered modelled *Mtb* transmission risk. Over the four weeks of implementation, these interventions were estimated to have prevented 16 infections.

Effective strategies for TB infection control in high-burden TB settings have been designated a research priority.^2^ We showed that simple IPC interventions targeting proximity between hospital attendees can effectively reduce *Mtb* transmission risk. While the redesigned waiting area and physical distancing measures reduced the spatial clustering among attendees and airborne *Mtb* transmission, the patient flow system added during the second intervention phase did not result in higher reductions than during the first intervention phase. Our local study team reported difficulties in enforcing the one-way movement pattern and it may have inadvertently increased local clustering on busy days by restricting access to certain areas. Nevertheless, the positive impact of the redesigned waiting area and physical distancing measures shows that low-cost infrastructure changes and simple cues can substantially lower the risk of short-range exposure and transmission.

Our findings align with modelling work from South Africa, which estimated that queue management systems incorporating outdoor waiting areas could substantially lower the rate of transmission within clinics.^3^ Similarly, our previous modelling showed that a package of IPC interventions during the COVID-19 pandemic, including an outdoor waiting area and restricted indoor patient flow, reduced the risk of *Mtb* transmission compared with pre-pandemic conditions.^6^ However, evidence on the real-world effectiveness of operational interventions to reduce crowding and proximity remains scarce. Our study provides empirical confirmation that such interventions can achieve a more even spatial distribution of patients, reducing short-range high particle–density exposure, thereby diminishing the contribution of close-contact to overall transmission risk.

Despite high patient volumes, the estimated risk of *Mtb* transmission in the Zambian hospital was low, probably owing to excellent natural ventilation. The median air exchange rate in the waiting area was 8 air changes per hour (ACH), exceeding the recommended standards for crowded indoor settings of 5 ACH by the US Centers for Disease Control and Prevention,^19^ and 6 ACH by The Lancet COVID-19 commission.^20^ In our spatiotemporal model, the air exchange rate influences both the diffusion and removal of infectious aerosols, thereby reducing transmission risk through dilution and replacement of contaminated air. We previously showed that if ventilation during the pandemic had been equivalent to pre-pandemic conditions, the risk of *Mtb* transmission would have more than doubled.^7^ This finding is consistent with data from a South African clinic, where enhanced natural ventilation through open doors and windows markedly reduced transmission,^21^ and from a Peruvian hospital, where structural modifications promoting airflow quadrupled the air exchange rate and lowered modelled infection risk by 72%.^22^

We detected *Mtb* DNA in the air of the hospital waiting room, demonstrating the possibility of transmission in our setting. Airborne transmission of *Mtb* was first demonstrated through classic experiments in which guinea pigs housed near tuberculosis wards became infected via shared air,^23^ providing foundational evidence for long-range aerosol spread. The Wells–Riley equation later quantified infection risk under the assumption of a well-mixed indoor airspace,^8^ in which infectious particles are evenly distributed. However, local airflow patterns and crowding can generate spatial heterogeneity, creating short-range “hotspots” of higher exposure that are not captured by well-mixed models.^24,25^ Local airflow patterns are difficult to measure in real-world settings, but in principle, empirical findings suggest that transmission risk of respiratory infections is inversely related to the distance to the infectious source.^26–28^ Our spatiotemporal model considered this by incorporating proximity and local aerosol diffusion, thereby enabling quantification of interventions that reduce crowding and short-range exposure.

Our study has several limitations. First, we assumed that people with confirmed TB and partly those with presumptive TB were infectious due to missed diagnosis, but the true distribution of infectiousness is unknown and asymptomatic individuals may also contribute to transmission.^29^ Second, we did not consider exposure outside the hospital’s waiting area. For example, attendees may have had additional exposure in consultation rooms or other areas of the hospital. Third, we could not empirically verify whether the modelled number of infections corresponded to the actual number of infections occurring in the hospital. However, *Mtb* transmission is difficult to measure due to the lack of an appropriate in vitro test assay,^30^ and to measure transmission as the number of secondary cases as determined by molecular epidemiology has its own limitations and is not feasible in the setting of a busy waiting room. Fourth, person-tracking data contained interruptions, preventing reliable estimation of individual-level infection risks; therefore, we report aggregate risks at the hospital level. Future studies integrating continuous tracking and detailed airflow measurements could help clarify how person-specific behaviour and microenvironmental variation influence TB transmission in clinical settings. Finally, although *Mtb* DNA was occasionally detected in bioaerosol samples from the waiting area, detections were rare—likely reflecting the hospital’s high ventilation rates as well as technical limitations in the molecular detection of *Mtb* in bioaerosol samples.

By integrating environmental, clinical, and person-movement data in a spatiotemporal model, our study showed how changes in the spatial clustering of people could lower short-range transmission of *Mtb*. Our findings showed that simple IPC interventions targeting indoor crowding and proximity can have a significant public health impact in reducing the risk of *Mtb* transmission in clinical settings. These results highlight the potential of low-cost infrastructure modifications and physical distancing measures as important components of infection prevention and control in high-burden, resource-limited health-care settings.

## Data Availability

All data (anonymous patient tracking, de-identified clinical records, and environmental measurements) will be made publicly available upon publication. Python and R code files used for data preprocessing and to perform transmission risk modelling and intervention effect estimation are publicly available via GitHub: https://github.com/nbanho/tb_patient_flows.git.

## Author contributions

NB and LuFe conceptualised and designed the study. NB and LuFe verified the data and analysed laboratory results from bioaerosol samples. NB conducted the formal investigation and performed the statistical and modelling analyses. GM, FM, EB and LuFe implemented the study and coordinated the interventions on site. PB and LaFu performed the laboratory analysis of bioaerosol samples. RS and DK contributed to data postprocessing. NB, ME, CB, and LuFe contributed to manuscript writing. All authors reviewed and approved the final version of the manuscript.

## Acknowledgements

This study was supported by the National Institute of Allergy and Infectious Diseases (NIAID) through grant no. 5U01-AI069924-05. NB is further supported by the Swiss National Science Foundation through grant no. P500PM_230473. The funders had no role in study design, data collection, data analysis, data interpretation, or writing of the report.

We are further grateful to the local healthcare workers and the hospital management staff for their involvement in designing the study and the interventions, and for their ongoing support throughout the study. We are indebted to the study staff at the hospital for their diligent daily work. Finally, we also would like to thank the IT staff at CIDRZ (Zambia) and ISPM (Switzerland), as well as ASE AG (Zürich, Switzerland), for their support with setting up the person tracking system and data transfer.

## Supplementary information

Supplementary Table S1. contains a detailed description of the implemented IPC interventions. Text A describes how interrupted person tracks were rejoined and person tracks of TB patients were identified using an R Shiny App, which is shown in Supplementary Figure S1. Text B describes how the air exchange rates were computed from intraday CO_2_ measurements (Supplementary Figure S2). Text C describes the spatiotemporal Wells-Riley model and the approach to estimate the effects of IPC interventions. Supplementary results are provided in Supplementary Tables S2–S3 and Figures S3-S11.

